# The Relationship Between Triglyceride to High-Density Lipoprotein Cholesterol Ratio and Cardiovascular High-Risk: A Cross-Sectional Investigation

**DOI:** 10.1101/2024.05.08.24307048

**Authors:** Pengcheng Li, Jirui Cai, Shuaifang Yuan, Yapeng Li, Huiting Shi, Cui Liang, Bing He, Qiaotao Xie, Baocang Lei, Jing Bai, Nan Wang, Dongliang Liu, Qichao Wang, Jianwei Xiong, Jin Wang, Haoran Wang

**Author notes:** Corresponding to: Haoran Wang, Luohe Central Hospital, Luohe, Medical College, Luohe, China.; Alternate corresponding author: Jin Wang, Luohe Central Hospital, Luohe Medical College, Luohe, China. Pengcheng Li and Jirui Cai contributed equally to this work.

## Abstract

**Aims:** The triglyceride to high-density lipoprotein cholesterol (TG/HDL-C) ratio reflects the balance between atherogenic and anti-atherogenic lipid fractions. The study aims to investigate the utility of the TG/HDL-C ratio in identifying high-risk groups for cardiovascular disease (CVD) within the general population.

**Methods:** The current study was a branch of the ChinaHEART cohort in middle China that involved a total of 6,593 community-dwelling adults. We examine the association between TG/HDL-C and CVD high-risk in a cross-sectional investigation.

**Results:** Results from restricted cubic spline and ROC analyses revealed a significant relationship between higher TG/HDL-C levels and increased odds of cardiovascular high-risk, with the ratio exhibiting good discriminatory power (AUC=0.819 in the fully adjusted model). Logistic regression further supported this association, indicating a 1.21-fold rise in the odds of high-risk for each unit increase in TG/HDL-C.

**Conclusions:** The TG/HDL ratio demonstrates potential as a valuable clinical tool for CVD prevention. By incorporating the TG/HDL ratio into risk assessment models, healthcare professionals can enhance risk stratification and identify high-risk individuals who may benefit from targeted interventions.

## Introduction

Cardiovascular diseases (CVD) continue to be a leading cause of mortality and morbidity worldwide, imposing a significant burden on healthcare systems and society as a whole. Early identification of individuals at high risk of developing CVD is crucial for implementing targeted prevention and management strategies.(2) Traditional risk factors such as hypertension, dyslipidemia, smoking, and diabetes have been widely recognized, but there is a growing interest in exploring novel biomarkers that may enhance risk stratification in the general population.(3)

In recent years, the triglyceride to high-density lipoprotein cholesterol (TG/HDL-C) ratio has emerged as a potential marker for cardiovascular risk assessment.(4–8) This ratio reflects the balance between atherogenic and anti-atherogenic lipid fractions. and has been proposed as a simple and cost-effective tool to identify individuals at increased risk of CVD. Some studies have found that this indicator is associated with morbidity and mortality from cardiovascular disease.(4,9) However, there is still a lack of relevant studies on whether this indicator can identify high-risk groups of cardiovascular disease in the general population.

Our study aims to address this gap in knowledge by conducting a cross-sectional observational study to evaluate the utility of the TG/HDL-C ratio in identifying high-risk individuals for cardiovascular diseases in the general population. By examining this novel biomarker in a large and diverse sample, we seek to elucidate its potential as a screening tool for early risk stratification and intervention. If we can effectively identify individuals at high risk of cardiovascular disease in the general population using this indicator, we can intervene at earlier stages of cardiovascular disease, thereby maximizing the prevention of major adverse cardiovascular events.

## Methods

### Study population and data collection

The current study is a branch of the China Health Evaluation And risk Reduction through nationwide Teamwork (ChinaHEART) in Luohe City, involving 6,859 community-dwelling adults recruited from two township hospitals in Luohe City between November 2, 2021 and February 20, 2022. The ChinaHEART cohort is a population cohort launched by the China National Center for Cardiovascular Diseases (NCCD) initially aiming at cardiovascular disease (CVD) risk screening and management.(10) Study methods has been described previously.(11) The study strictly adhered to ethical guidelines, with approval granted by the Ethics Committee of Fuwai Hospital and the Ethics Committee of Luohe Central Hospital. Participants received detailed information on the study’s purpose, significance, and risks before participant, in accordance with the Declaration of Helsinki. Written informed consent was obtained from all participants. The inclusion criteria for this study are as follows: (1) Age: 35-75 years (born between January 1, 1946, and December 31, 1986); (2) Residing near the survey site for at least 6 months in the 12 months prior to screening; (3) Voluntarily participating in this project and signing the informed consent form. The exclusion criteria are as follows: (1) Survey participants with missing data; (2) Survey participants who answered “unclear” to questionnaire items; (3) Survey participants with fasting time less than 8 hours before blood collection.

We utilized standardized questionnaires to collect data on socio-demographic characteristics (e.g., age, gender), lifestyle factors (e.g., smoking, alcohol consumption), medical history (including hypertension, diabetes, dyslipidemia, coronary heart disease, myocardial infarction, stroke), and medication usage (such as antihypertensive, hypoglycemic, and lipid-lowering drugs) of the study participants. Data were collected through an information system provided by the ChinaHEART study and was uploaded directly to the national study center in the Fuwai Hospital. The anonymous data used in the current study was applied from the national study center in the Fuwai Hospital after they performed data washing. Cardiovascular disease (CVD) was defined as a diagnosis of coronary heart disease, previous myocardial infarction, or stroke. Body mass index (BMI) was calculated as body mass in kilograms divided by the square of height in meters. Waist circumference was measured by placing a measuring tape horizontally around the abdomen at the midpoint between the anterior superior iliac crest and the lower edge of the twelfth rib. Systolic blood pressure (SBP) and diastolic blood pressure (DBP) were measured in a seated resting position at least twice, and the average values were recorded. Lipid profiles were assessed using fingertip blood samples.

According to the risk assessment prediction map in the Guidelines for Cardiovascular Risk Assessment and Management issued by the World Health Organization in 2008, the risk assessment of cardiovascular disease was conducted.(12) If the screening subjects had 10 years of risk of cardiovascular disease 20%, they were judged as high risk.

### Statistics

The study population was categorized based on quartiles of the TG/HDL-C ratio for basic characteristic description. Normality of continuous variables was assessed using the Kolmogorov-Smirnov test, with mean ± standard deviation for normally distributed data, and median (25%-75% interval) for non-normally distributed data. Categorical variables were described using percentages. Distribution differences in high cardiovascular risk proportions across TG/HDL-C ratio quartile populations were illustrated with a percentage stacking bar graph. Continuous variable comparisons between groups involved t-tests or analysis of variance for normally distributed data, and rank sum tests for non-normally distributed data. Chi-square tests were conducted for categorical variable comparisons. The association between the TG/HDL-C ratio and cardiovascular high risk was examined using logistic regression analysis, with two models utilized: one without confounder adjustments for univariate analysis, and another adjusting for confounding factors including gender, age, marital status, BMI, waist circumference, fasting blood glucose, smoking history, alcohol consumption history, diabetes, and systolic and diastolic blood pressure. Nonlinear associations between the TG/HDL-C ratio and cardiovascular risk were explored through restricted cubic spline analysis, utilizing key points at the 10th, 25th, 50th, 75th, and 90th percentiles of the TG/HDL-C ratio. The ability of the TG/HDL-C ratio to identify high cardiovascular risk was evaluated using receiver operating characteristic (ROC) curve analysis. While conducting the ROC curve analysis, we calculated the appropriate cutoff value, and this cutoff value was used to divide the participants into the high and low TG/HDL-C groups for further logistic regression analysis. Furthermore, we used net reclassification improvement (NRI) index to quantify the added value of TG/HDL-C for the discriminative performance.

## Results

A total of 6,593 participants with sufficient data were included in the study, comprising 1,417 individuals categorized as cardiovascular high-risk and 5,176 individuals in the non-high-risk category. The study cohort was stratified into four segments based on the TG/HDL-C ratio, highlighting notable variations in demographic and clinical characteristics such as age, gender, height, weight, BMI, waist circumference, smoking habits, alcohol consumption, history of hypertension, diabetes, as well as levels of glucose, lipids, and blood pressure (refer to Table 1). The distribution of high-risk individuals across these quartiles was 16%, 15%, 18%, and 38% respectively (view Figure 1).

**Figure 1.**
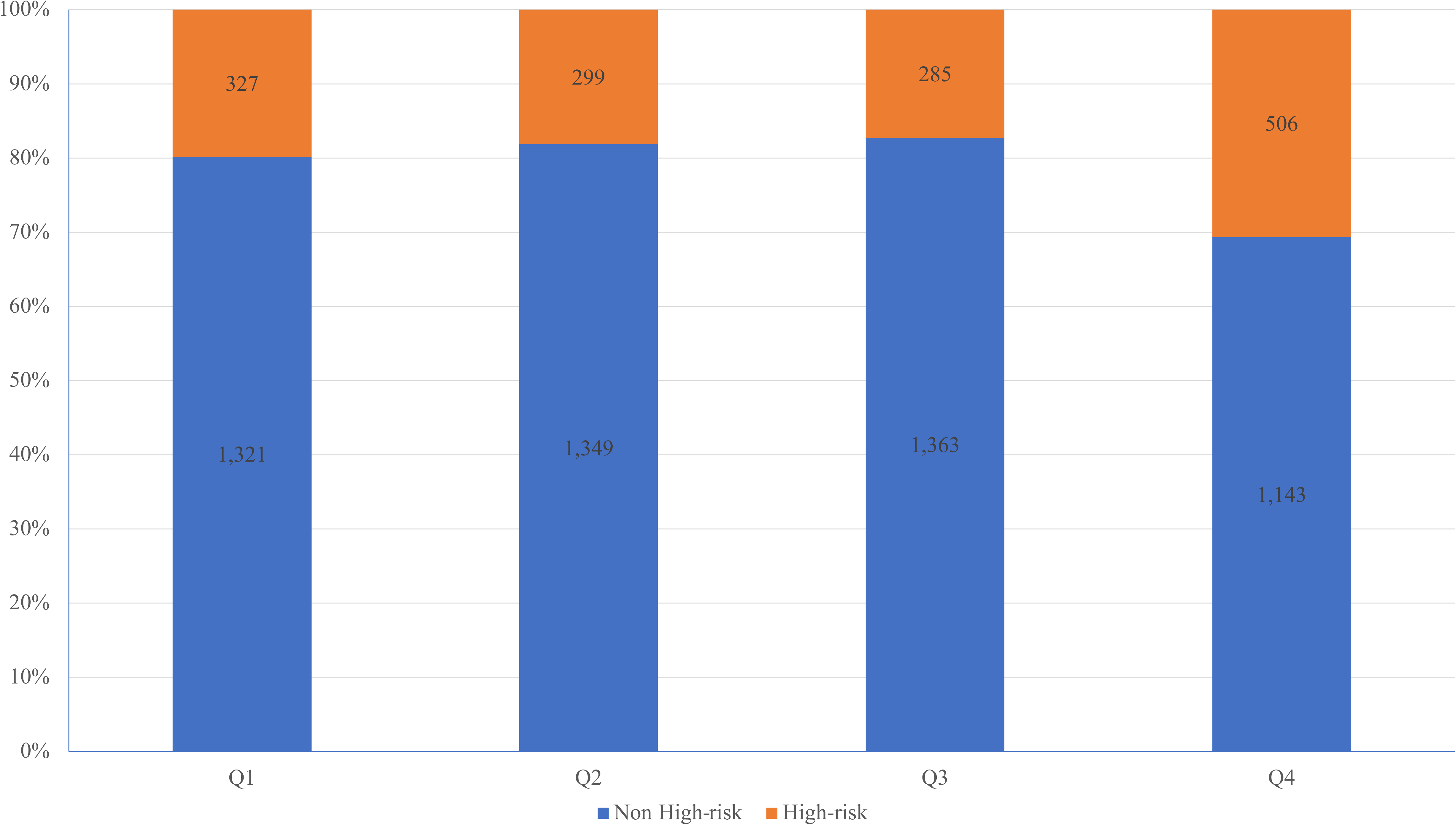
Percentage stacked histogram to visually represent the distribution of cardiovascular high-risk individuals across quartiles of the TG/HDL-C ratio.

**Table 1.**
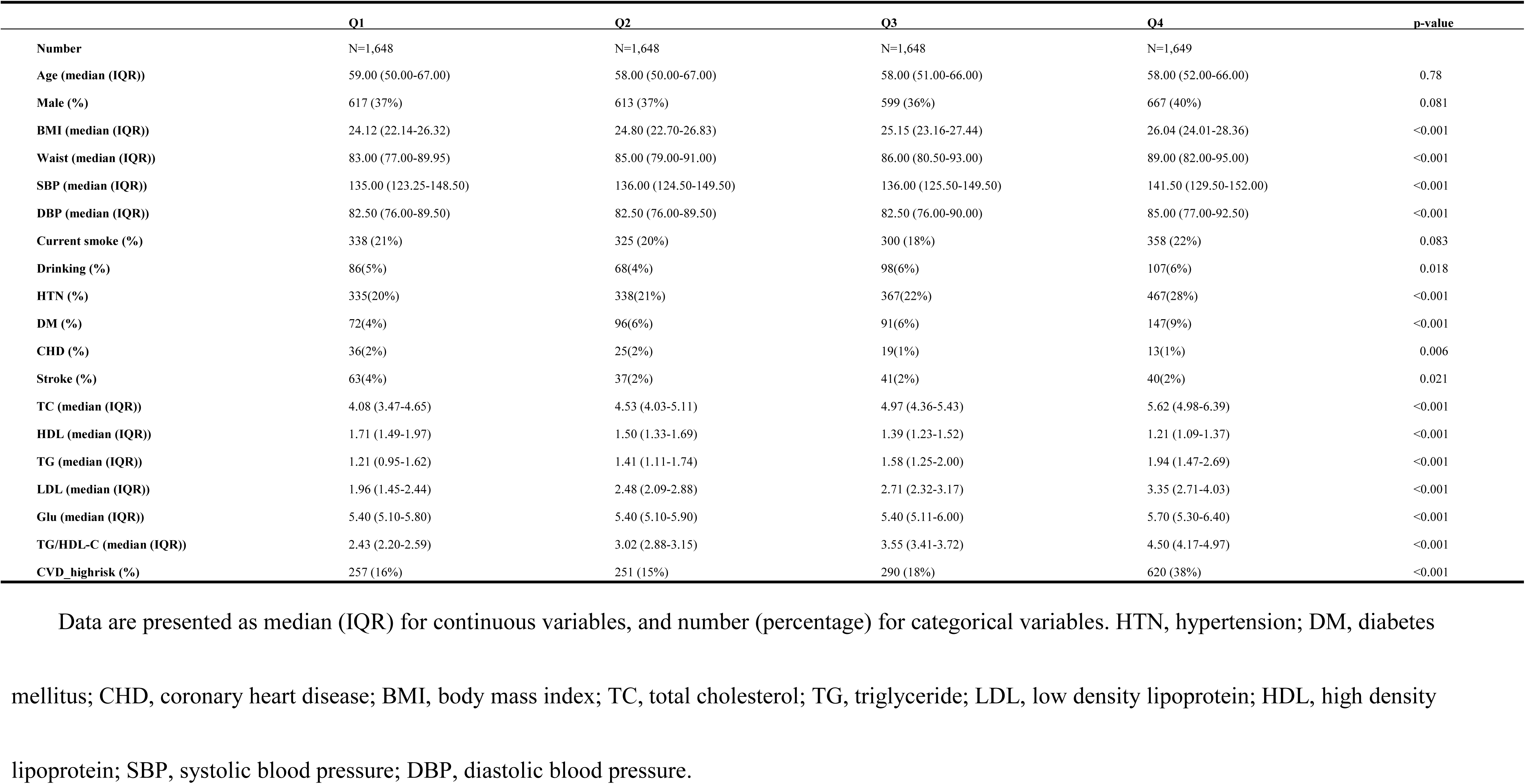
Clinical characteristics of the study population by quartiles of TG/HDL-C.

|  | Q1 | Q2 | Q3 | Q4 | p-value |
| --- | --- | --- | --- | --- | --- |
| **Number** | N=1,648 | N=1,648 | N=1,648 | N=1,649 |  |
| **Age (median (IQR))** | 59.00 (50.00-67.00) | 58.00 (50.00-67.00) | 58.00 (51.00-66.00) | 58.00 (52.00-66.00) | 0.78 |
| **Male (%)** | 617 (37%) | 613 (37%) | 599 (36%) | 667 (40%) | 0.081 |
| **BMI (median (IQR))** | 24.12 (22.14-26.32) | 24.80 (22.70-26.83) | 25.15 (23.16-27.44) | 26.04 (24.01-28.36) | <0.001 |
| **Waist (median (IQR))** | 83.00 (77.00-89.95) | 85.00 (79.00-91.00) | 86.00 (80.50-93.00) | 89.00 (82.00-95.00) | <0.001 |
| **SBP (median (IQR))** | 135.00 (123.25-148.50) | 136.00 (124.50-149.50) | 136.00 (125.50-149.50) | 141.50 (129.50-152.00) | <0.001 |
| **DBP (median (IQR))** | 82.50 (76.00-89.50) | 82.50 (76.00-89.50) | 82.50 (76.00-90.00) | 85.00 (77.00-92.50) | <0.001 |
| **Current smoke (%)** | 338 (21%) | 325 (20%) | 300 (18%) | 358 (22%) | 0.083 |
| **Drinking (%)** | 86(5%) | 68(4%) | 98(6%) | 107(6%) | 0.018 |
| **HTN (%)** | 335(20%) | 338(21%) | 367(22%) | 467(28%) | <0.001 |
| **DM (%)** | 72(4%) | 96(6%) | 91(6%) | 147(9%) | <0.001 |
| **CHD (%)** | 36(2%) | 25(2%) | 19(1%) | 13(1%) | 0.006 |
| **Stroke (%)** | 63(4%) | 37(2%) | 41(2%) | 40(2%) | 0.021 |
| **TC (median (IQR))** | 4.08 (3.47-4.65) | 4.53 (4.03-5.11) | 4.97 (4.36-5.43) | 5.62 (4.98-6.39) | <0.001 |
| **HDL (median (IQR))** | 1.71 (1.49-1.97) | 1.50 (1.33-1.69) | 1.39 (1.23-1.52) | 1.21 (1.09-1.37) | <0.001 |
| **TG (median (IQR))** | 1.21 (0.95-1.62) | 1.41 (1.11-1.74) | 1.58 (1.25-2.00) | 1.94 (1.47-2.69) | <0.001 |
| **LDL (median (IQR))** | 1.96 (1.45-2.44) | 2.48 (2.09-2.88) | 2.71 (2.32-3.17) | 3.35 (2.71-4.03) | <0.001 |
| **Glu (median (IQR))** | 5.40 (5.10-5.80) | 5.40 (5.10-5.90) | 5.40 (5.11-6.00) | 5.70 (5.30-6.40) | <0.001 |
| **TG/HDL-C (median (IQR))** | 2.43 (2.20-2.59) | 3.02 (2.88-3.15) | 3.55 (3.41-3.72) | 4.50 (4.17-4.97) | <0.001 |
| **CVD_highrisk (%)** | 257 (16%) | 251 (15%) | 290 (18%) | 620 (38%) | <0.001 |

Data are presented as median (IQR) for continuous variables, and number (percentage) for categorical variables. HTN, hypertension; DM, diabetes
mellitus; CHD, coronary heart disease; BMI, body mass index; TC, total cholesterol; TG, triglyceride; LDL, low density lipoprotein; HDL, high density
lipoprotein; SBP, systolic blood pressure; DBP, diastolic blood pressure.

To explore the nonlinear relationship between the TG/HDL-C ratio and cardiovascular high-risk, a restricted cubic spline analysis was conducted. The analysis revealed a positive association between elevated TG/HDL-C ratio levels and an increased likelihood of cardiovascular high-risk (see Figure 2).

**Figure 2.**
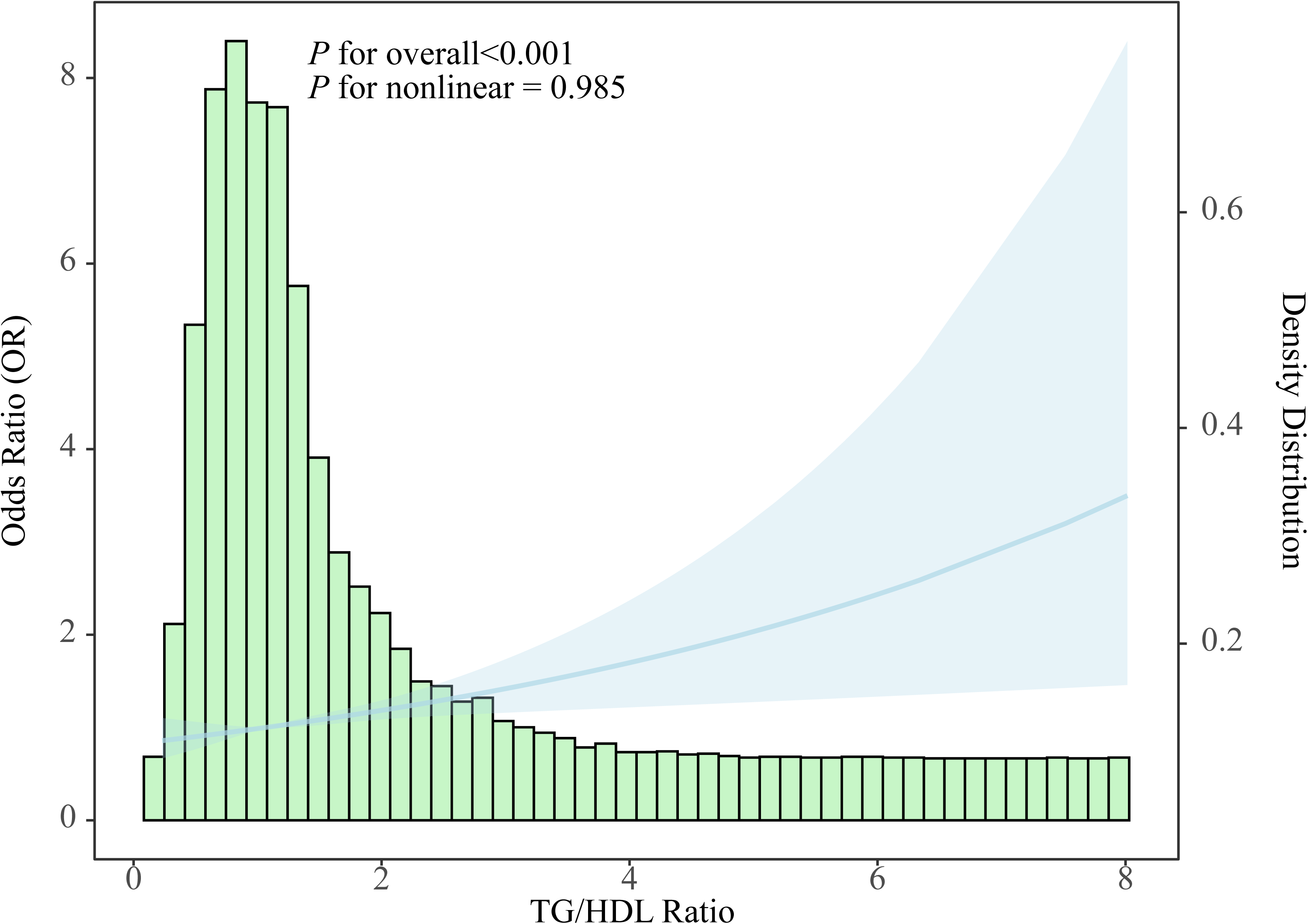
Distribution histogram of the TG/HDL-C ratio in the study population and restricted cubic spline to assess the relationship between TG/HDL-C ratio and cardiovascular high-risk.

The TG/HDL-C ratio’s discriminatory ability in identifying individuals at cardiovascular high-risk was evaluated through ROC analysis (refer to Figure 3). Initially, in model 1, the AUC for the TG/HDL-C ratio alone was 0.562 (95% CI 0.544-0.580) for all participants. However, upon incorporating all covariates into the analysis in model 2, the AUC significantly increased to 0.819 (95% CI 0.805-0.834), suggesting the enhanced effectiveness of combining the TG/HDL-C ratio with traditional cardiovascular risk factors for high-risk identification. Notably, the NRI index showed a significant improvement in discriminative performance, with a value of 0.107 ± 0.030 (p = 0.016). Moreover, the ROC analysis identified the optimal cutoff value for the TG/HDL-C ratio as 1.18, enabling the classification of participants into high and low TG/HDL-C ratio groups.

**Figure 3.**
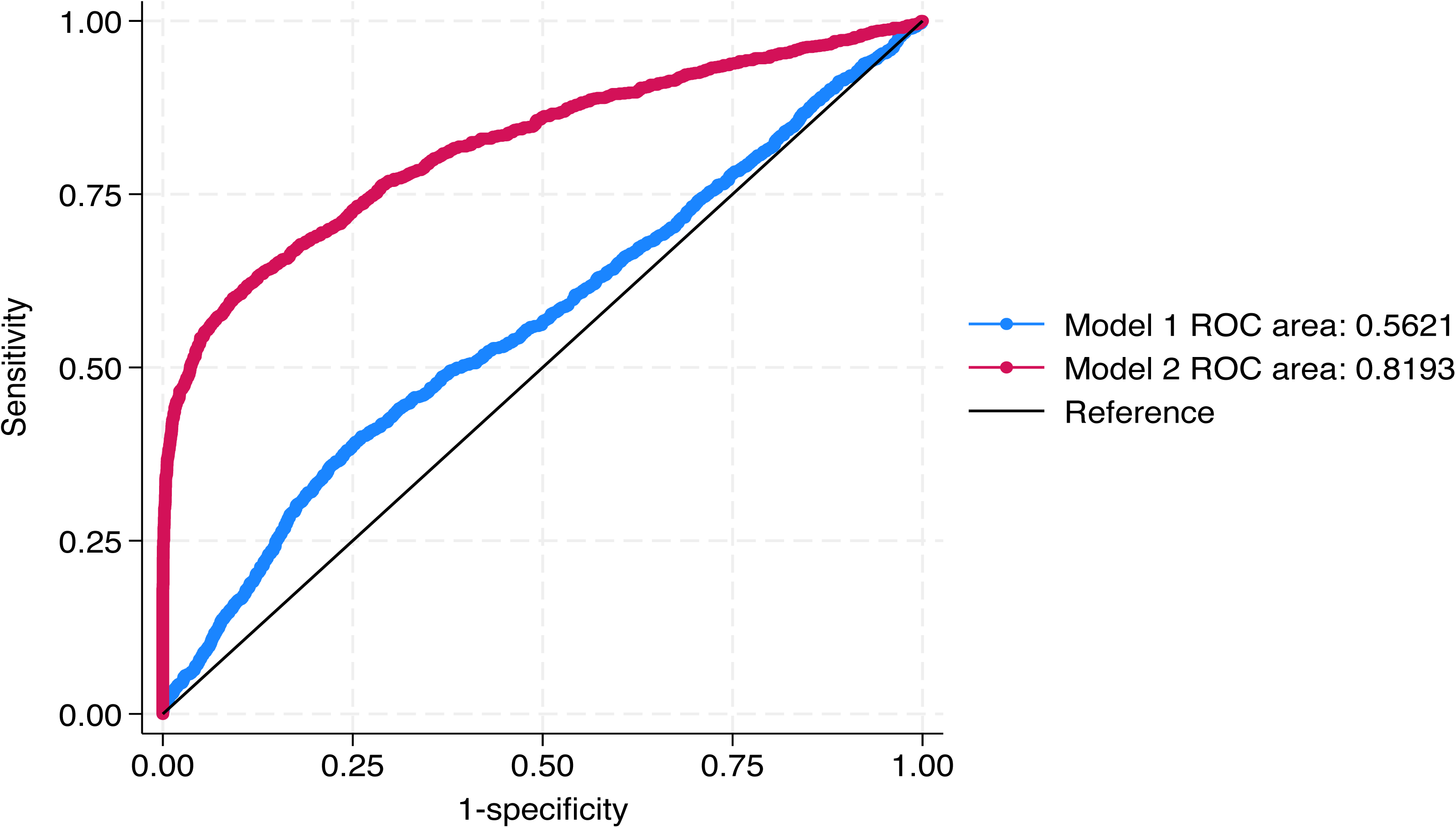
ROC curve to illustrate the discriminatory power of TG/HDL-C ratio for cardiovascular high-risk.

The logistic regression analysis results examining the relationship between the TG/HDL-C ratio and cardiovascular high-risk are detailed in Table 2. In Model 1, a 1 unit increase in the TG/HDL-C ratio was associated with a 1.40-fold increase in the odds of cardiovascular high-risk (95% CI 1.30-1.51, p < 0.001). Comparatively, the high TG/HDL-C ratio group exhibited 1.61 times higher odds of high-risk than the low TG/HDL-C ratio group (95% CI 1.43-1.81, p < 0.001), while the highest quartile of the TG/HDL-C ratio had 1.79 times higher odds compared to the lowest quartile (95% CI 1.52-2.10, p < 0.001). In Model 2, each 1 unit rise in the TG/HDL-C ratio was associated with a 1.21-fold increase in the odds of cardiovascular high-risk (95% CI 1.11-1.32, p < 0.001). Additionally, the high TG/HDL-C ratio group demonstrated 1.27 times higher odds of high-risk compared to the low TG/HDL-C ratio group (95% CI 1.10-1.47, p = 0.001), with the highest quartile of the TG/HDL-C ratio displaying 1.49 times higher odds than the lowest quartile (95% CI 1.21-1.83, p < 0.001).

**Table 2.** Association between TG/HDL-C ratio and cardiovascular high-risk by logistic regression.

|  | Model 1 |  | Model 2 |  |
| --- | --- | --- | --- | --- |
|  | OR (95% CI) | p | OR (95% CI) | p |
| **All participants** |  |  |  |  |
| TG/HDL-C (per 1 unit) | 1.40 (1.30-1.51) | <0.001 | 1.21 (1.11-1.32) | <0.001 |
| TG/HDL-C (high vs low) | 1.61 (1.43-1.81) | <0.001 | 1.27 (1.10-1.47) | 0.001 |
| TG/HDL-C (quartile) |  |  |  |  |
| Q1 | Ref |  | Ref |  |
| Q2 | 0.90 (0.75-1.07) | 0.214 | 1.04 (0.85-1.29) | 0.68 |
| Q3 | 0.84 (0.71-1.01) | 0.06 | 0.95 (0.77-1.18) | 0.634 |
| Q4 | 1.79 (1.52-2.10) | <0.001 | 1.49 (1.21-1.83) | <0.001 |
| **Female** |  |  |  |  |
| TG/HDL-C (per 1 unit) | 1.36 (1.24-1.50) | <0.001 | 1.14 (1.01-1.28) | 0.028 |
| TG/HDL-C (high vs low) | 1.52 (1.30-1.77) | <0.001 | 1.16 (0.96-1.40) | 0.114 |
| TG/HDL-C (quartile) |  |  |  |  |
| Q1 | Ref |  | Ref |  |
| Q2 | 0.94 (0.74-1.19) | 0.6 | 0.97 (0.74-1.28) | 0.838 |
| Q3 | 0.91 (0.72-1.15) | 0.447 | 1.00 (0.76-1.32) | 0.998 |
| Q4 | 1.78 (1.44-2.20) | <0.001 | 1.33 (1.02-1.74) | 0.034 |
| **Male** |  |  |  |  |
| TG/HDL-C (per 1 unit) | 1.50 (1.33-1.69) | <0.001 | 1.31 (1.14-1.52) | <0.001 |
| TG/HDL-C (high vs low) | 1.82 (1.51-2.19) | <0.001 | 1.45 (1.15-1.84) | <0.001 |
| TG/HDL-C (quartile) |  |  |  |  |
| Q1 | Ref |  | Ref |  |
| Q2 | 0.85 (0.66-1.11) | 0.238 | 1.14 (0.84-1.57) | 0.432 |
| Q3 | 0.78 (0.59-1.03) | 0.076 | 0.90 (0.64-1.27) | 0.557 |
| Q4 | 1.94 (1.51-2.49) | <0.001 | 1.67 (1.20-2.32) | 0.002 |

## Discussion

Cardiovascular disease (CVD) remains a significant global health burden, necessitating the identification of individuals at high risk for effective prevention and intervention strategies. The TG/HDL-C ratio has emerged as a potential marker for assessing cardiovascular risk in the general population. It has been observed that the ratio of TG/HDL-C ratio not only provides an approximation of insulin resistance but also assists in identifying patients with an atherogenic lipoprotein profile.(13) The typical lipoprotein phenotype in insulin-resistant/hyperinsulinemic patients is typified by high plasma TG and low HDL cholesterol levels, smaller and denser LDL particles, as well as an increased accumulation of remnant lipoproteins after a meal. This lipoprotein phenotype is not only associated with insulin resistance, but it is also linked with an elevated risk of cardiovascular disease.(14) Although plasma TG and HDL cholesterol levels are commonly tested, the same cannot be said for the LDL particle diameter or postprandial remnant concentrations. Nonetheless, a previous study observed a robust correlation between the TG/HDL cholesterol ratio and the LDL peak diameter.(15) Moreover, it is suggested that a TG/HDL cholesterol concentration ratio of 3.5 can accurately predict the presence of the small dense LDL phenotype (LDL phenotype B). Therefore, by assessing plasma TG and HDL cholesterol levels and computing their ratio, it is possible to obtain understanding into three out of the four modifications in lipoprotein metabolism that enhance the risk of cardiovascular disease in insulin-resistant patients.

Numerous epidemiological studies have examined the relationship between the TG/HDL ratio and CVD risk. Most of these studies have reported a positive association between a higher TG/HDL ratio and increased CVD risk. For instance, a prospective cohort study by Gaziano et al. found the ratio of triglycerides to HDL was a strong predictor of myocardial infarction;(16) Urbina et al. suggests the TG/HDL ratio may be a useful marker for identifying young individuals at risk for developing arterial stiffness, an early sign of CVD;(17) Li et al. found a positive correlation between the TG/HDL-C ratio and carotid intima thickness, a marker of early atherosclerosis, in adolescents with type 2 diabetes;(18) Du et al. found that the TG/HDL ratio is associated with insulin resistance, a major risk factor for CVD, in a Chinese population;(19) Ren et al. found that the TG/HDL ratio is positively associated with insulin resistance in newly diagnosed type 2 diabetes patients, indicating an increased risk of CVD;(20) Squillace et al. found that a higher TG/HDL ratio is predictive of the development of diabetes, a major risk factor for CVD;(21) Hong et al. found that a high TG/HDL ratio is associated with an increased risk of cardiovascular events in patients with diabetes and coronary artery disease.(22)

The TG/HDL ratio holds clinical significance as a potential marker for CVD prevention. It provides valuable insights into the balance between TG and HDL levels, representing the interplay between dyslipidemia and atherosclerosis. A higher TG/HDL ratio reflects an increased atherogenic lipid profile and has been associated with arterial stiffness, carotid intima thickness, insulin resistance and diabetes, all of which contribute to CVD development. By incorporating the TG/HDL ratio into risk assessment models, clinicians can more accurately identify individuals at high risk for CVD and implement targeted interventions.

Despite the clinical significance of the TG/HDL ratio, several limitations must be acknowledged. First, the cut-off values for defining high TG/HDL ratios vary across studies, leading to heterogeneity in research findings. Standardization of these thresholds is necessary to ensure consistency and comparability between studies. Additionally, confounding factors, including age, sex, lifestyle factors, and comorbidities, may influence the association between the TG/HDL ratio and CVD risk. Although some studies have attempted to account for these factors, further research is needed to elucidate their precise impact. Moreover, variations in laboratory methods for lipid profile assessment may introduce measurement errors and affect the accuracy of TG and HDL cholesterol estimations.

In conclusion, the TG/HDL ratio demonstrates potential as a valuable clinical tool for CVD prevention. Its ability to capture the interplay between triglycerides and HDL cholesterol provides a comprehensive assessment of the atherogenic lipid profile. By incorporating the TG/HDL ratio into risk assessment models, healthcare professionals can enhance risk stratification and identify high-risk individuals who may benefit from targeted interventions. However, standardization of cut-off values, better control of confounding factors, and harmonization of laboratory methods are needed to further validate the clinical significance of the TG/HDL ratio. With these advancements, the TG/HDL ratio could significantly contribute to the prevention and control of cardiovascular disease.

## Data Availability

The data underlying the results presented in the study are available from Haoran Wang.

## Availability of data and materials

The data that support the findings of this study are available from the corresponding author Haoran Wang, upon reasonable request.

## Author Contributions statement

Conceptualization and Methodology: Haoran Wang, Pengcheng Li; Formal analysis: Pengcheng Li, Jirui Cai, Shuaifang Yuan, Yapeng Li, Huiting Shi, Cui Liang, Bing He; Data Curation: Qiaotao Xie, Baocang Lei,Jing Bai, Nan Wang, Dongliang Liu, Qichao Wang, Jianwei Xiong, Jin Wang, Haoran Wang; Writing: Pengcheng Li, Jirui Cai, Shuaifang Yuan, Yapeng Li, Huiting Shi, Cui Liang; Funding acquisition: Qiaotao Xie, Haoran Wang, Pengcheng Li.

## Ethical statement

This study was conducted with approval from the Ethics Committee of Fuwai Hospital (2014–574) as the Central Ethic, and was approved by the Ethics Committee of Luohe Central Hospital as the local ethic. This study was conducted in accordance with the declaration of Helsinki. Written informed consent was obtained from all participants.

